# Association Between Insulin Therapy and Cardiovascular Morbidity in Adults with Type 2 Diabetes: Analysis of BRFSS 2022–2023 Data

**DOI:** 10.1101/2025.08.07.25333260

**Authors:** Ahmad Assinnari, Salman Althobaiti

## Abstract

**Background:** Insulin therapy is a cornerstone in the management of Type 2 Diabetes Mellitus (T2DM), although its relationship with cardiovascular outcomes remains an area of discussion. While it effectively used in managing either types of diabetes, evidence regarding its cardiovascular safety is mixed.

**Objective:** To investigate the association between insulin therapy, timing and duration, and cardiovascular outcomes among adults with T2DM, while accounting for biological, psychological, and social factors, using data from the 2022–2023 Behavioral Risk Factor Surveillance System (BRFSS).

**Methods:** Multivariate logistic regression was used on a de-identified cross-sectional BRFSS data of adults with self-reported T2DM to estimate the association between insulin use and cardiovascular outcomes, while adjusting for age, hypertension, dyslipidemia, smoking status, psychological comorbidities, and medication use.

**Results:** Insulin users exhibited significantly higher rates of coronary heart disease (21.8% vs. 14.2%) and stroke (13.4% vs. 6.6%) compared to non-users (p < 0.001). It was found that early initiation and longer duration of insulin therapy were independently associated with increased cardiovascular risk. These associations persisted after adjusting for biological and psychosocial variables.

**Conclusion:** In this nationally representative sample, insulin therapy, particularly when initiated early or sustained long-term, was associated with higher odds of cardiovascular morbidity in adults with T2DM. These findings highlight the need for individualized treatment approaches that consider both glycemic control and broader biopsychosocial factors.

## Introduction

The therapeutic landscape of Type 2 Diabetes Mellitus (T2DM) has undergone notable evolution, yet insulin therapy has retained its central role in achieving glycemic regulation and preventing long-term metabolic complications. Despite this, the clinical positioning of insulin remains contested, particularly regarding its association with cardiovascular outcomes, which still constitute the primary burden of morbidity and mortality among individuals with T2DM [1]. Such ambiguity calls for a closer inspection not only of insulin’s biochemical effects but also of the clinical logic behind its initiation and sustained use across diverse patient trajectories.

Cardiovascular disease (CVD) is the leading cause of death among individuals with T2DM, highlighting the importance of refining therapeutic approaches to reduce this risk [2]. The relationship between insulin administration and cardiovascular outcomes is multifaceted, influenced by factors such as the patient’s age at insulin initiation and the total duration of therapy. Some studies suggest that early insulin administration may delay the progression of cardiovascular disease by improving glycemic control and mitigating ‘glucotoxicity’ [2,3]. However, other research has reported potential adverse outcomes associated with insulin therapy, depending on treatment regimens and individual metabolic responses [3,4]. Several large observational studies and meta-analyses have identified increased risks of cardiovascular events and all-cause mortality among insulin-treated patients, particularly those with existing cardiovascular conditions or heart failure [5,6]. These risks may be mediated by insulin-related hypoglycemia, fluid retention, weight gain, or inflammatory effects [5,6]. Conversely, randomized controlled trials such as ORIGIN have shown neutral results, while early intensive insulin therapy in newly diagnosed patients has occasionally demonstrated protective effects against stroke and heart failure [7]. The DIGAMI trial reported a post-MI mortality reduction, though this was not replicated in DIGAMI-2 [8,9]. These discrepancies may reflect differences in study populations, glycemic targets, and background therapies.

What complicates the picture further is insulin’s biological footprint beyond glycemia. Its role in modulating lipid metabolism, endothelial function, and inflammatory signaling is increasingly recognized [10]. These non-glycemic pathways intersect meaningfully with cardiovascular risk and may mediate the observed clinical outcomes in ways that have yet to be fully modeled. Hence, the therapeutic implications of insulin are inseparable from the patient’s broader metabolic and cardiovascular milieu.

Given the conflicting evidence, the present study aims to contribute to this ongoing discussion by examining how the timing and duration of insulin therapy are associated with cardiovascular outcomes in a nationally representative sample of adults with T2DM. We further explore how biological, psychological, and social factors may interact with insulin use in shaping cardiovascular risk. Data were obtained from the 2022–2023 BRFSS survey.

## Methodology

### Data Collection

This cross-sectional study analyzed de-identified data from the Behavioral Risk Factor Surveillance System (BRFSS) for the years 2022 and 2023, focusing on adults with self-reported Type 2 Diabetes Mellitus (T2DM). To ensure a uniform study population representative of sustained insulin therapy, individuals with Type 1 Diabetes and those who had discontinued insulin use were excluded. Survey-specific sampling weights provided by BRFSS were applied to account for the complex sampling design, enhancing the representativeness and generalizability of findings to the broader U.S. T2DM population.

### Data Analysis

All statistical analyses were conducted using SPSS software, version 27 (IBM Corp., Armonk, NY, USA). Preliminary statistical analyses utilized Chi-square tests to evaluate associations between insulin utilization and categorical variables, including hypertension and dyslipidemia. Continuous variables such as age and duration of insulin use were examined using independent samples T-tests to compare means among various patient groups. Additionally, logistic regression analyses were utilized to determine predictors of cardiovascular outcomes. Univariate models evaluated the effects of singular variables, whereas multivariate models accounted for confounding factors including age, hypertension, antihypertensive medication use, anxiety, depression, dyslipidemia, smoking status (both traditional and electronic cigarettes), utilization of diabetic medications, and current insulin therapy. A p-value below 0.05 was deemed statistically significant for all analyses. Cramér’s V was employed to quantify the strength of associations, while Cohen’s d was utilized to measure the magnitude of differences.

## Results

### Demographic and Clinical Characteristics

Table 1. provides a comprehensive overview of the baseline demographic and clinical characteristics of the study cohorts. Insulin users had a mean age of 62.57 ± 13.30 years, which was slightly younger than non-insulin users at 63.8 ± 12.68 years, with a statistically significant difference (p < 0.001). The percentage of males was slightly elevated among insulin users, suggesting a potential gender-related predisposition or management strategy in insulin therapy. The incidence of hypertension and the administration of antihypertensive medications were markedly elevated among insulin users, indicating a more intricate clinical profile with a potentially increased cardiovascular risk burden (78.8% vs. 74.2% for hypertension, p < 0.001; 93.8% vs. 90.8% for antihypertensive use, p < 0.001).

**Table 1.** Demographic and Clinical Characteristics by Treatment Group.

|  | Total | Insulin users | Non-Insulin users | P-value |  |
| --- | --- | --- | --- | --- | --- |
| **Age (mean ± SD)** | 62.89±13.14 | 63.8 ± 12.68 | 62.57 ± 13.30 | <0.001 | |
| Gender (male, %) | 20143393 | 5275123<br>(49.6) | 14868270<br>(50.4) | <0.001 | 0.01 |
| Hypertension (yes, %) | 30278171(75.4) | 8383035<br>(78.8) | 21895136<br>(74.2) | <0.001 | 0.05 |
| Antihypertensive medications (yes, %) | 27707930 (91.6) | 7852255(93.8) | 19855675<br>(90.8) | <0.001 | 0.05 |
| Anxiety (yes, %) | 8471083 (21.1) | 2660456 (25) | 5810627 (19.7) | <0.001 | 0.06 |
| Depression (yes, %) | 10281470 (25.5) | 3430081(32.2) | 6851389 (23.2) | <0.001 | 0.09 |
| Dyslipidemia (yes, %) | 26405010 (66) | 7365862<br>(69.6) | 19039148<br>(64.6) | <0.001 | 0.05 |
| Conventional smoking (yes, %) | 16991076 (43.5) | 4534086<br>(44.1) | 12456990<br>(43.3) | <0.001 | 0.01 |
| Electronic cigarettes smoking (yes, %) | 4571012 (11.7) | 1153198<br>(11.2) | 3417814 (11.9) | <0.001 | 0.01 |
| Coronary heart disease (yes, %) | 6480484 (16.2) | 2311733<br>(21.8) | 4168751 (14.2) | <0.001 | 0.10 |
| Angina (yes, %) | 2281123 (5.7) | 895482 (8.4) | 1385641 (4.7) | <0.001 | 0.71 |
| MI (yes, %) | 4180491(10.4) | 1575847<br>(14.8) | 2604644 (8.8) | <0.001 | 0.09 |
| Stroke (yes, %) | 3382753 (8.4) | 1423199<br>(13.4) | 1959554 (6.6) | <0.001 | 0.11 |
| Diabetic pills (yes, %) | 31516687 (78.5) | 7256589<br>(68.2) | 24260098<br>(82.3) | <0.001 | 0.15 |

### Cardiovascular Outcomes

**Table 2** emphasizes the significant differences in the incidence of cardiovascular complications between insulin users and non-insulin users. Insulin users exhibited a markedly elevated prevalence of coronary heart disease (21.8% vs. 14.2%), myocardial infarction (14.8% vs. 8.8%), stroke (13.4% vs. 6.6%), and angina (8.4% vs. 4.7%), all with p-values < 0.001. These findings indicate an increased cardiovascular risk linked to insulin therapy that requires further investigation and meticulous management.

**Table 2.**
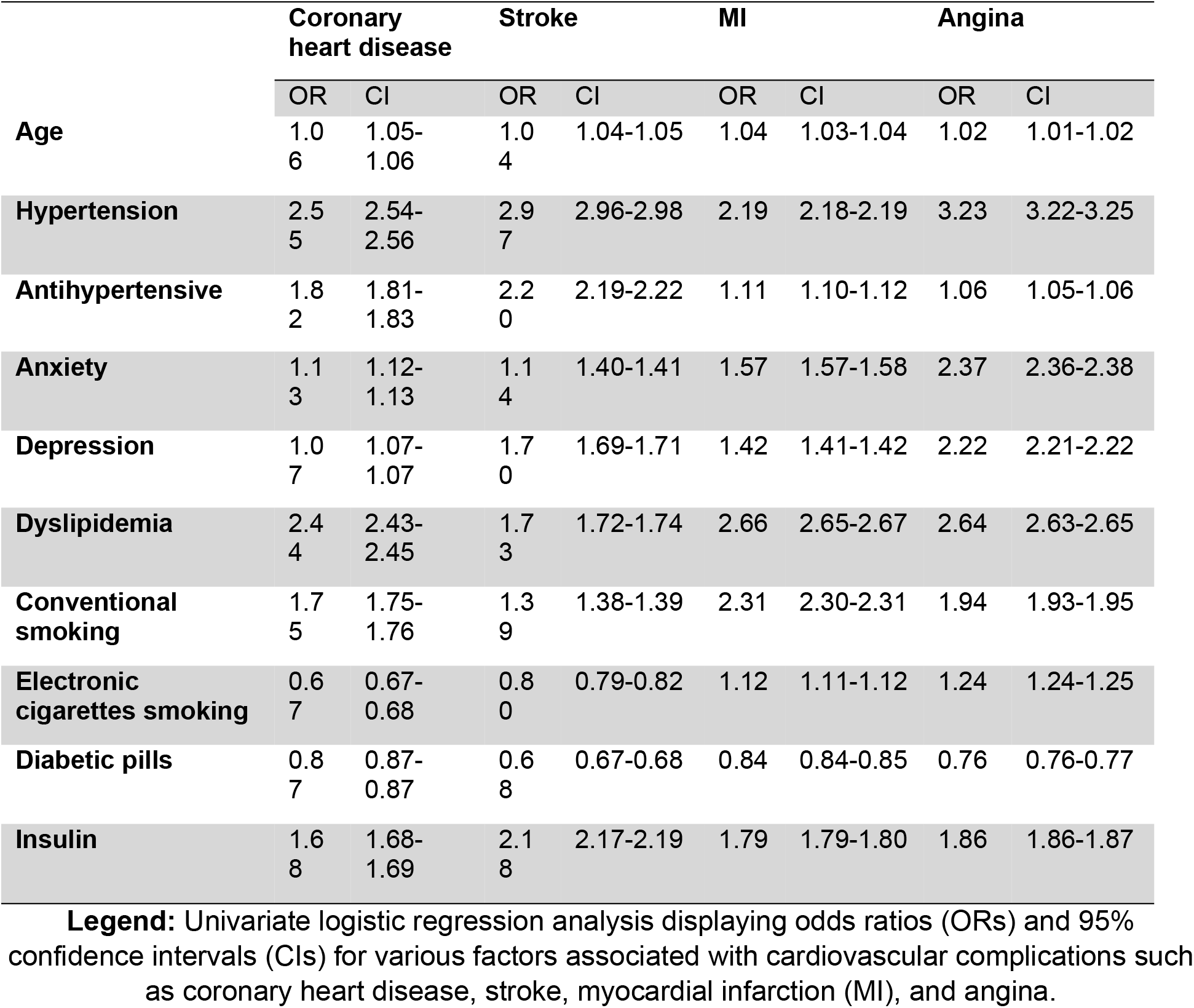
Association of Clinical Factors with Cardiovascular Outcomes in T2DM.

### Logistic Regression Analysis

**Table 3** presents findings from the multivariate logistic regression analysis, accounting for various confounding variables. The sustained presence of markedly increased odds ratios for major cardiovascular events among insulin users is clinically relevant. After accounting for potential confounders such as hypertension, dyslipidemia, and smoking, insulin utilization was linked to a significant elevation in the risk of coronary heart disease (OR = 1.52, 95% CI 1.52-1.53) and stroke (OR = 1.93, 95% CI 1.93-1.94). This indicates an inherent connection between insulin therapy and cardiovascular morbidity that cannot be entirely elucidated by other prevalent risk factors.

**Table 3.**
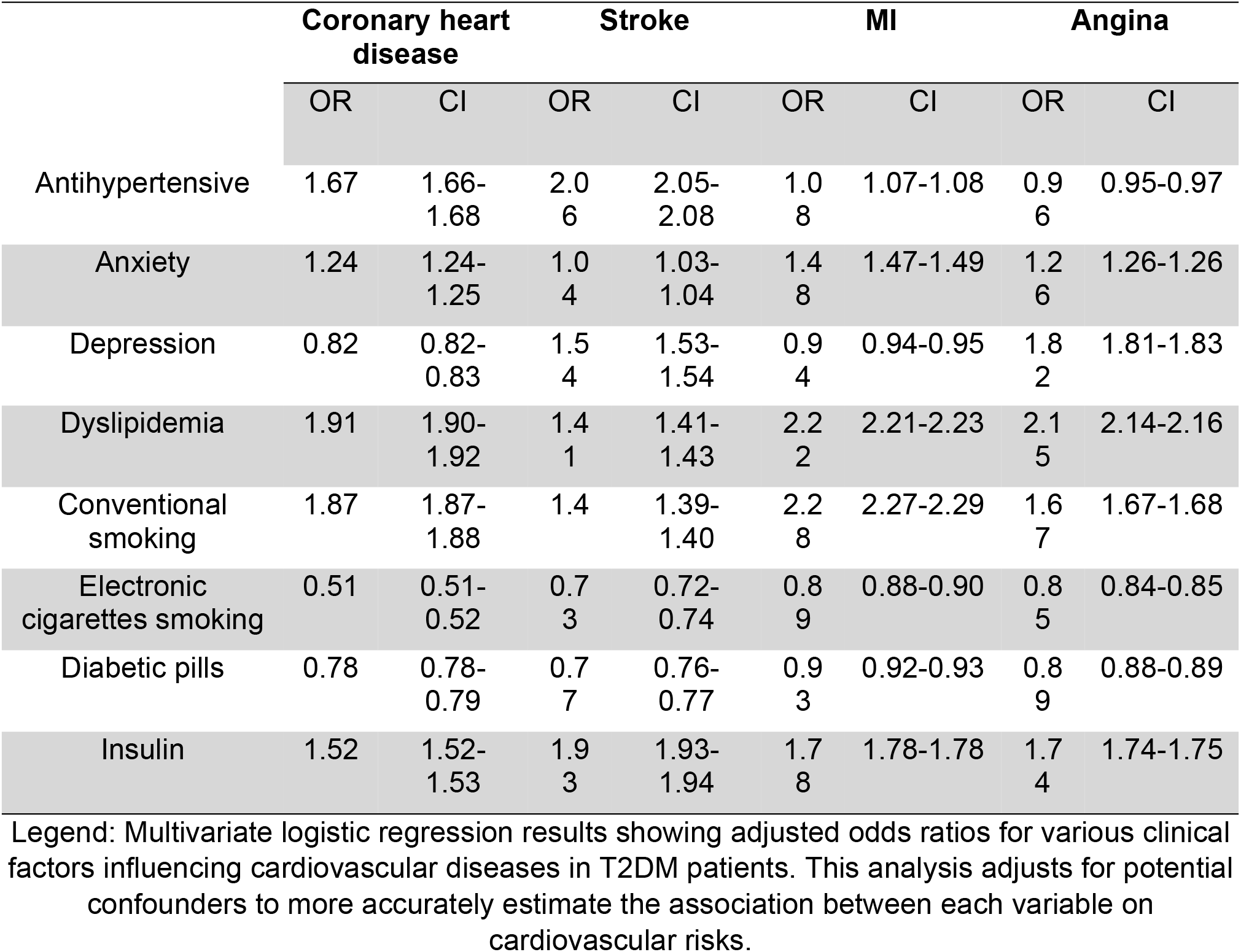
Adjusted Associations Between Clinical Factors and Cardiovascular Outcomes.

### Impact of Insulin Therapy Dynamics

**Table 4** explores the dynamics of insulin therapy, analyzing the influence of timing and duration on cardiovascular risk. Initiating insulin therapy within the inaugural year of type 2 diabetes mellitus diagnosis was associated with an elevated risk of coronary heart disease (OR = 1.11, 95% CI 1.10-1.11) and myocardial infarction (OR = 1.21, 95% CI 1.21-1.22). Moreover, the length of insulin administration was associated with heightened cardiovascular risks, particularly for myocardial infarction (OR = 1.02 per annum, 95% CI 1.02-1.02) and stroke (OR = 1.00 per annum, 95% CI 1.00-1.00). These findings highlight the essential requirement for strategic timing and meticulous oversight of insulin therapy to reduce potential risks.

**Table 4.**
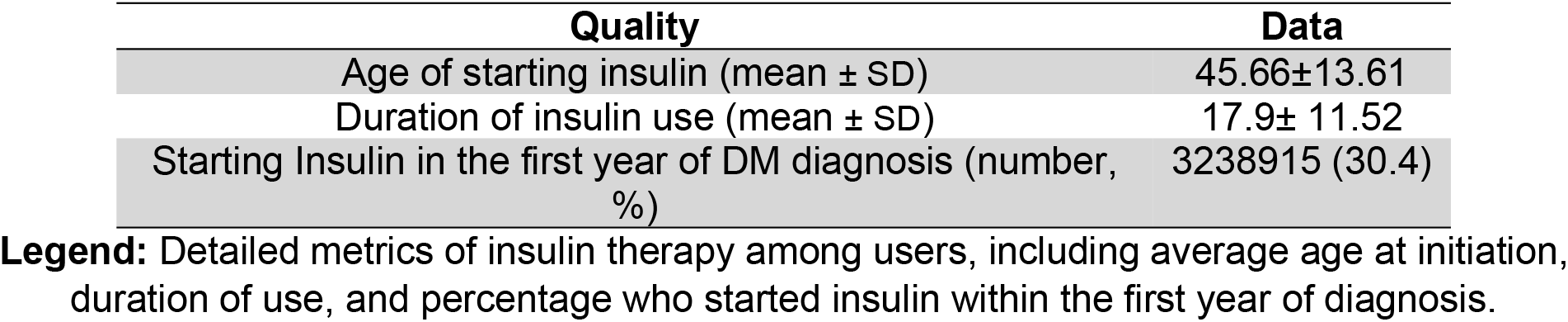
Characteristics of Insulin Therapy Among Users.

| Quality | Data |
| --- | --- |
| Age of starting insulin (mean ± SD) | 45.66±13.61 |
| Duration of insulin use (mean ± SD) | 17.9± 11.52 |
| Starting Insulin in the first year of DM diagnosis (number, %) | 3238915 (30.4) |

**Legend:** Detailed metrics of insulin therapy among users, including average age at initiation, duration of use, and percentage who started insulin within the first year of diagnosis.

## Discussion

This study investigates the association between insulin therapy and cardiovascular complications and various health metrics in patients with T2DM, providing significant insights into its relationship with age at onset and duration of insulin administration. Our findings underscore a notable correlation between insulin utilization and cardiovascular disease (CVD), specifically identifying higher prevalence of coronary heart disease (CHD), myocardial infarction (MI), and stroke among insulin users relative to non-insulin users. Given insulin’s known role in glycemic control, these associations raise important clinical considerations. Our analysis reveals that the length of insulin therapy is associated with an increased likelihood of adverse cardiovascular events, consistent with the findings of Luo et al., which indicated that early insulin therapy did not significantly diminish the risk of coronary heart disease but was linked to a reduced incidence of stroke and heart failure hospitalization in newly diagnosed T2DM patients.[11]

The comparison of insulin therapy with other antidiabetic treatments, especially SGLT2 inhibitors, indicates varying cardiovascular outcomes. SGLT2 inhibitors demonstrate protective advantages against cardiovascular disease in patients with type 2 diabetes mellitus, in contrast to the neutral or adverse cardiovascular outcomes linked to insulin. [12] This aligns with the cardioprotective effects of SGLT2 inhibitors on heart failure and mortality noted in larger cohort.[13] An essential element of insulin therapy is its correlation with hypoglycemia, which has been associated with cardiovascular incidents and mortality. Our findings indicate that a nuanced understanding is essential, as the cardiovascular risks associated with insulin may be mediated by hypoglycemic episodes, especially in patients with prolonged diabetes or considerable comorbidities. [13] This is further supported by studies indicating that hypoglycemia can trigger arrhythmic events, compounding cardiovascular risk. [13]

The significance of insulin resistance as a determinant of cardiovascular risk in insulin-treated patients is paramount. Patients exhibiting elevated insulin resistance demonstrate an increased risk profile for cardiovascular disease events while undergoing insulin therapy. [14] This relationship underscores the need for personalized treatment approaches that consider the underlying metabolic status of the patient.

Recent studies have highlighted the intricacies of managing cardiovascular risks in patients newly diagnosed with T2DM who are commencing insulin therapy. Lee et al. indicate that insulin therapy in newly diagnosed T2DM patients was associated with a higher risk of cardiovascular events and overall mortality compared to non-diabetic individuals, underscoring the need for rigorous cardiovascular protective measures in this population.[15]

Jeon et al. elucidate the varying effects of early insulin initiation, indicating a greater risk of microvascular complications associated with insulin compared to oral antidiabetic medications, while no heightened risks for macrovascular complications or overall mortality were detected, implying the complex implications of insulin therapy on long-term diabetes outcomes.[16]

Currie et al. identified a U-shaped correlation between HbA1c levels and mortality in patients with type 2 diabetes, indicating that both low and high HbA1c levels correlate with elevated risks, with the minimum hazard occurring at a HbA1c of approximately 7.5%.[17] This finding aligns with Colayco et al., who found increased cardiovascular events at both extremely low and high A1C levels relative to an intermediate range, underscoring the risks of both under-treatment and over-treatment in the diabetic population. [18] Choi et al. studied the potential increase of oxidative stress by acute hyperglycemia, which may contribute to the adverse effects observed at both low and high glucose levels, suggesting that glucose variability could be a pivotal element in diabetes management. [19] Collectively, these studies highlight the intricacy of glycemic regulation in diabetes, indicating that sustaining moderate, stable glucose levels may reduce risk and prevent the negative consequences linked to the extremes of glycemic control.

Recent studies have examined the complex role of insulin therapy and its broader health implications beyond glycemic regulation. The research conducted by Zhu et al. offers a thorough analysis of insulin resistance indices and their relationship with all-cause mortality in individuals with diabetic kidney disease, indicating that particular levels of insulin resistance are markedly linked to mortality risks, highlighting the intricate role of insulin in the advancement of chronic diseases.[20] Fazio et al. investigate the systemic ramifications of hyperinsulinemia, associating persistently elevated insulin levels with an augmented risk of type 2 diabetes, cardiovascular diseases, as well as heightened occurrences of cancer and neurodegenerative disorders. [21] In a notable study, Singh et al. examine the effects of insulin therapy on cardiovascular outcomes in patients with T2DM, specifically addressing the cardiovascular risks linked to varying durations and intensities of insulin administration. Their findings indicate that intensive insulin therapy, especially in individuals with chronic diabetes, might increase cardiovascular risk, thus endorsing a measured approach to insulin delivery. [22] Additionally, the research by Parikh et al provides valuable insights into the genetic factors that may influence insulin resistance and its subsequent impact on the efficacy of insulin therapy. Their study suggests that genetic predispositions can significantly affect insulin sensitivity, thereby influencing the overall effectiveness and safety of insulin therapy in managing diabetes.[23]

## Data Availability

All relevant data are within the manuscript and its Supporting Information files.

https://www.cdc.gov/brfss/annual_data/annual_2023.html
.

## Limitations and Future Directions

While our study provides substantial evidence on the complex interactions between insulin therapy and cardiovascular health, several limitations must be acknowledged. The observational nature of our data limits causal inferences.

Additionally, the heterogeneity in patient health status and treatment regimens suggests that further randomized controlled trials are needed to delineate the specific conditions under which insulin therapy may be beneficial or harmful.

## Conclusion

Our findings suggest that insulin therapy, particularly when administered without careful consideration of individual patient factors, such as insulin resistance and duration of diabetes, may be associated with increased cardiovascular risk. This emphasizes the importance of a personalized approach to diabetes management, prioritizing newer therapeutic options that confer cardiovascular protection in appropriate patients.

## Data Availability Statement

The data used in this study are publicly available through the Centre of Disease Control (CDC) website: https://www.cdc.gov/brfss/annual_data/annual_2023.html.

## Ethics Statement

The data that were used in the study are de-identified, publicly available secondary data from BRFSS. Therefore, no ethical approval under institutional or national guidelines is required.

## References

1 Ma CX, Ma XN, Guan CH, Li YD, Mauricio D, Fu SB. Cardiovascular disease in type 2 diabetes mellitus: progress toward personalized management. Cardiovasc Diabetol. 2022;21(1):74. Published 2022 May 14. doi:10.1186/s12933-022-01516-6

2 Lu J, Wang C, Shen Y, et al. Time in Range in Relation to All-Cause and Cardiovascular Mortality in Patients With Type 2 Diabetes: A Prospective Cohort Study. Diabetes Care. 2021;44(2):549–555. doi:10.2337/dc20-1862

3 Yang Y, Cai Z, Zhang J. Insulin Treatment May Increase Adverse Outcomes in Patients With COVID-19 and Diabetes: A Systematic Review and Meta-Analysis. Front Endocrinol (Lausanne). 2021;12:696087. Published 2021 Jul 22. doi:10.3389/fendo.2021.696087

4 Yu B, Li C, Sun Y, Wang DW. Insulin Treatment Is Associated with Increased Mortality in Patients with COVID-19 and Type 2 Diabetes. Cell Metab. 2021;33(1):65-77.e2. doi:10.1016/j.cmet.2020.11.014

5 Herman ME, O’Keefe JH, Bell DSH, Schwartz SS. Insulin Therapy Increases Cardiovascular Risk in Type 2 Diabetes. Prog Cardiovasc Dis. 2017;60(3):422–434. doi:10.1016/j.pcad.2017.09.001

6 Liu J, Hu X. Impact of insulin therapy on outcomes of diabetic patients with heart failure: A systematic review and meta-analysis. Diab Vasc Dis Res. 2022;19(3):14791641221093175. doi:10.1177/14791641221093175

7 ORIGIN Trial Investigators, Gerstein HC, Bosch J, et al. Basal insulin and cardiovascular and other outcomes in dysglycemia. N Engl J Med. 2012;367(4):319–328. doi:10.1056/NEJMoa1203858

8 Malmberg K, Rydén L, Efendic S, et al. Randomized trial of insulin-glucose infusion followed by subcutaneous insulin treatment in diabetic patients with acute myocardial infarction (DIGAMI study): effects on mortality at 1 year. J Am Coll Cardiol. 1995;26(1):57–65. doi:10.1016/0735-1097(95)00126-k

9 Malmberg K, Rydén L, Wedel H, et al. Intense metabolic control by means of insulin in patients with diabetes mellitus and acute myocardial infarction (DIGAMI 2): effects on mortality and morbidity. Eur Heart J. 2005;26(7):650–661. doi:10.1093/eurheartj/ehi199

10 Marušić M, Paić M, Knobloch M, Liberati Pršo AM. NAFLD, Insulin Resistance, and Diabetes Mellitus Type 2. Can J Gastroenterol Hepatol. 2021;2021:6613827. Published 2021 Feb 17. doi:10.1155/2021/6613827

11 Luo S, Zheng X, Bao W, et al. Real-world effectiveness of early insulin therapy on the incidence of cardiovascular events in newly diagnosed type 2 diabetes. Signal Transduct Target Ther. 2024;9(1):154. Published 2024 Jun 6. doi:10.1038/s41392-024-01854-9

12 Wang W, Chen LY, Walker RF, et al. SGLT2 Inhibitors Are Associated With Reduced Cardiovascular Disease in Patients With Type 2 Diabetes: An Analysis of Real-World Data. Mayo Clin Proc. 2023;98(7):985–996. doi:10.1016/j.mayocp.2023.01.023

13 Andersen A, Jørgensen PG, Knop FK, Vilsbøll T. Hypoglycaemia and cardiac arrhythmias in diabetes. Ther Adv Endocrinol Metab. 2020;11:2042018820911803. Published 2020 May 19. doi:10.1177/2042018820911803

14 Mendez CE, Walker RJ, Eiler CR, Mishriky BM, Egede LE. Insulin therapy in patients with type 2 diabetes and high insulin resistance is associated with increased risk of complications and mortality. Postgrad Med. 2019;131(6):376–382. doi:10.1080/00325481.2019.1643635

15 Lee YB, Han K, Kim B, et al. Risk of early mortality and cardiovascular disease according to the presence of recently diagnosed diabetes and requirement for insulin treatment: A nationwide study. J Diabetes Investig. 2021;12(10):1855–1863. doi:10.1111/jdi.13539

16 Jeon HL, Kim W, Kim B, Shin JY. Relationship between the early initiation of insulin treatment and diabetic complications in patients newly diagnosed with type 2 diabetes mellitus in Korea: A nationwide cohort study. J Diabetes Investig. 2022;13(5):830–838. doi:10.1111/jdi.13719

17 Currie CJ, Peters JR, Tynan A, et al. Survival as a function of HbA(1c) in people with type 2 diabetes: a retrospective cohort study. Lancet. 2010;375(9713):481–489. doi:10.1016/S0140-6736(09)61969-3

18 Colayco DC, Niu F, McCombs JS, Cheetham TC. A1C and cardiovascular outcomes in type 2 diabetes: a nested case-control study. Diabetes Care. 2011;34(1):77–83. doi:10.2337/dc10-1318

19 Choi SW, Benzie IF, Ma SW, Strain JJ, Hannigan BM. Acute hyperglycemia and oxidative stress: direct cause and effect?. Free Radic Biol Med. 2008;44(7):1217–1231. doi:10.1016/j.freeradbiomed.2007.12.005

20 Low CY, Gan WL, Lai SJ, et al. Critical updates on oral insulin drug delivery systems for type 2 diabetes mellitus. J Nanobiotechnology. 2025;23(1):16. Published 2025 Jan 15. doi:10.1186/s12951-024-03062-7

21 Loktionova MV, Mohammadian M, Choopani R, Kheiri S, Mohammadian-Hafshejani A. Investigating the relationship between insulin use and all-cause mortality, breast cancer mortality, and recurrence risk in diabetic patients with breast cancer: A comprehensive systematic review and meta-analysis. PLoS One. 2024;19(12):e0314565. Published 2024 Dec 5. doi:10.1371/journal.pone.0314565

22 S Y, S V,T T J, et al. Understanding the Complexity of Hyperglycemic Emergencies: Exploring the Influence of the Type and Duration of Diabetes Mellitus and Its Impact on Mortality. Cureus. 2024;16(4):e58916. Published 2024 Apr 24. doi:10.7759/cureus.58916

23 Parikh HM, Elgzyri T, Alibegovic A, et al. Relationship between insulin sensitivity and gene expression in human skeletal muscle. BMC Endocr Disord. 2021;21(1):32. Published 2021 Feb 27. doi:10.1186/s12902-021-00687-9

